# Contradictory task demands among junior physicians during post-graduate training: An explorative, multi-methods study

**DOI:** 10.1101/2025.07.21.25331908

**Authors:** Matthias Weigl, Susann Böhm, Caroline Quartucci

## Abstract

**Introduction:** Junior physicians often face high levels of work stress, burnout, and depression, largely due to challenging work conditions. Contradictory task demands – a specific job stressor where junior physicians are expected to fulfill conflicting goals simultaneously - has received limited attention, but may significantly impact their mental health and job satisfaction.

**Objective:** This study aimed to identify key contradictory demands faced by junior physicians in specialty training and examine associations with physician health and well-being.

**Methods:** Step-wise and multi-methods procedure comprising explorative interviews and standardized surveys. A convenience sample of junior physicians undergoing specialty training in Germany were surveyed. Thematic analyses of qualitative data as well as descriptive and bivariate analyses were performed.

**Results:** Drawing upon seven in-depth interviews, experiences of key contradictory demands were extracted and consolidated into 19 distinct statements. 28 Survey responses were obtained with highest ratings for ‘Non-physician activities, additional tasks versus Focus on core clinical tasks’ as well as ‘General workload versus Regular working time’. Associations with physicians’ health and well-being outcomes were moderate and inconsistent.

**Discussion:** Preliminary results shed light on experiences of contradictory task demands in clinical work among junior physicians. A broad spectrum of potentially incompatible or conflicting demands on the job was observed. The findings underscore the complexity of the clinical work environment, where multiple conflicting demands create a particularly challenging educational and work setting for junior physicians. The results may inform tailored approaches to mitigate work stress in this professional group.

## INTRODUCTION

Junior physicians undergoing specialty education and post-graduate training repeatedly report significant work stress and mental health burden: several studies within in this professional group found high prevalence of work stress with particular high rates of burnout and depression (1-4). Foremost, this high proportion of mental health burden amongst junior physicians has been attributed to difficult work conditions during first years of clinical training and work practice (5). Workload and work environment factors have been repeatedly found to influence experiences of work stress and mental health burden in junior physicians (6-8).

A specific work stressor in hospitals, that has received less attention is the issue of ‘contradictory task demands’. We define contradictory task demands, if requirements are imposed that interfere with each other or are incompatible to complete without additional efforts. This has also been termed as contradictory job requirements (9). Especially in care work environments, junior physicians can come into conflict if they are required to attain multiple goals than cannot be aligned smoothly, i.e., efficient patient care, obtaining new skills, complete specialty curriculum on time.

Eventually, contradictory requirements make it challenging to address all demands adequately or urge junior physicians to identify priorities (9-11). Experiences of contradictory demands may eventually lead to difficulties in task performance, increased stress, or reduced efficiency (10). This conflict can arise from differing objectives stemming from the work environment, organizational, time constraints or other limited and inadequate resources (9, 12). For example, in clinical work, patient-related stress and overall time pressure may trigger different responses and strategies to attend those diverging demands, i.e., spending more time with difficult patient cases versus overall time resources within a constrained clinical work day (13).

Despite the relevance of contradictory task demands in junior physicians’ work life, respective evidence and literature base is still limited. Amidst the broad scope of adverse work conditions that may affect junior physicians’ mental health, deeper insights into potentially contradictory task demands imposed through the clinical work environment are missing (5). There is particularly lack of insights into the scope of potential contradictory demands during specialty training, especially in multiple-goal work environments (14). Junior physicians in specialty training cope with a challenging transition period from medical training in university into clinical practice, where often already high levels of mental health burden stemming from intense academic education are observed and that may be sustained into post-graduate specialty training (15). Although this phenomenon of contradictory task demands has been well described conceptually (9) and qualitatively (16), specific measures such as scales or item pools to assess this particular stressor in clinical work environments are missing. Lastly, conflicting task demands are triggering work stress experiences since available resources and different demands are not matching (9, 17). Yet, associations of contradictory demands with health care provider outcomes are sparse (16, 17). We therefore sought to explore relationships of experiences of contradictory demands with junior physicians’ well-being outcomes, such as affective (i.e., burnout, depressive symptoms) as well as cognitive outcomes (i.e., job satisfaction, intention to leave). Poor well-being on the job may affect safety and quality of patient care including poor patient satisfaction or interpersonal aspects of care (18, 19).

## Objectives

In the light of the above mentioned shortcomings, our exploratory investigation amongst junior physicians undergoing post-graduate specialty training sought to explore two aims:

1. to identify key contradictory demands and classification into major domains (step 1), and
2. to explore associations with physician health and well-being outcomes (step 2).

## METHODS

### Design

A consecutive, step-wise, multi-methods investigation was established: First, non-standardized exploratory interviews with junior physicians to obtain in-depth information concerning conflicting task demands in clinical practice (step 1) and a standardized survey (step 2). Prior to start, the study protocol was approved by the Ethics committee of the Medical Faculty of Ludwig-Maximilians-University Munich (No 17-305). For comprehensive reporting of our study, we used the GRAMMS recommendations, Good Reporting of A Mixed Methods Study (20)

### Sample

For step 1, we sought to explore reports and experiences of conflict task demands among junior physicians. Eligibility criteria were (1) physicians undergoing postgraduate specialty education (i.e., with maximum 5 years after graduation), (2) capable of German language, (3) working in direct patient care in Germany, (4) with no major mental or physical health issues. For step 2, again, we surveyed junior physicians with the identical eligibility criteria as in previous step 1.

### Procedure

For step 1, a convenience sampling approach was undertaken through the study team. Using a snowball sampling technique, information on the study was distributed through the personal network of the study team. After providing informed consent, each interview was conducted in person with one member of the study team and audio-taped. All information gathered, were anonymized. Duration of interviews ranged between 15 and 30 Minutes.

For step 2, again, a convenience sample of junior physicians was surveyed. Participants were recruited through the personal networks of the study authors and mailing lists among junior physicians in South Germany. The survey was sent to participants who expressed interest together with study information, consent form, and an additional, pre-validated return envelope.

### Measures

For step 1, the exploratory interviews, we used open-ended questions for exploring experiences and situation of conflicting task demands, defined as demands in medical practice that are perceived as incompatible. We further asked participants to prioritize dominant, frequent situations of conflicting task demands in their daily clinical practice. Before start, we pre-defined four distinct domains where conflicting demands may occur:

1. (I Structural demands) Demands imposed by and requirements rooted in the organization and employer: Requirements placed on physicians by superiors/employers, clinic operators, the healthcare system or society.
2. (II Patient-related demands) … by patients or through direct patient interaction
3. (III Individual demands) … in subjectively important goals and subjective needs of the providers (i.e., subjectively significant professional goals in life).
4. (IV Extra-professional demands) … placed on the physician outside of the profession or by her/his private life

After providing a short definition for each domain, interviewees were asked to answer the following questions respectively: (1) to report experiences of conflicting demands in the respective domain, (2) to explain and comment in detail on the reports. At the end, interviewees were asked to add any further issues or information relevant to the study question. Before use, a cognitive pre-test of the interview guidelines and questions were conducted with a non-involved junior physician.

For step 2 (quantitative survey), a standardized, paper-based survey was handed out. First, participants were provided with a short definition of conflicting demands. Then a list of 19 potentially relevant conflicting demands (i.e., developed in step 1) was presented. Each conflicting demand was presented as an items formulated as an antipode of two incompatible demands, (for the full list of surveyed items, see Table 1). All participants were required to report on the frequency of experiences per each statement. Scale range (1 ‘never’, 2 ‘very rarely’, 3 ‘rather rarely’, 4 ‘occasionally’, 5 ‘rather frequent’, 6 ‘very frequent’).

**Table 1:**
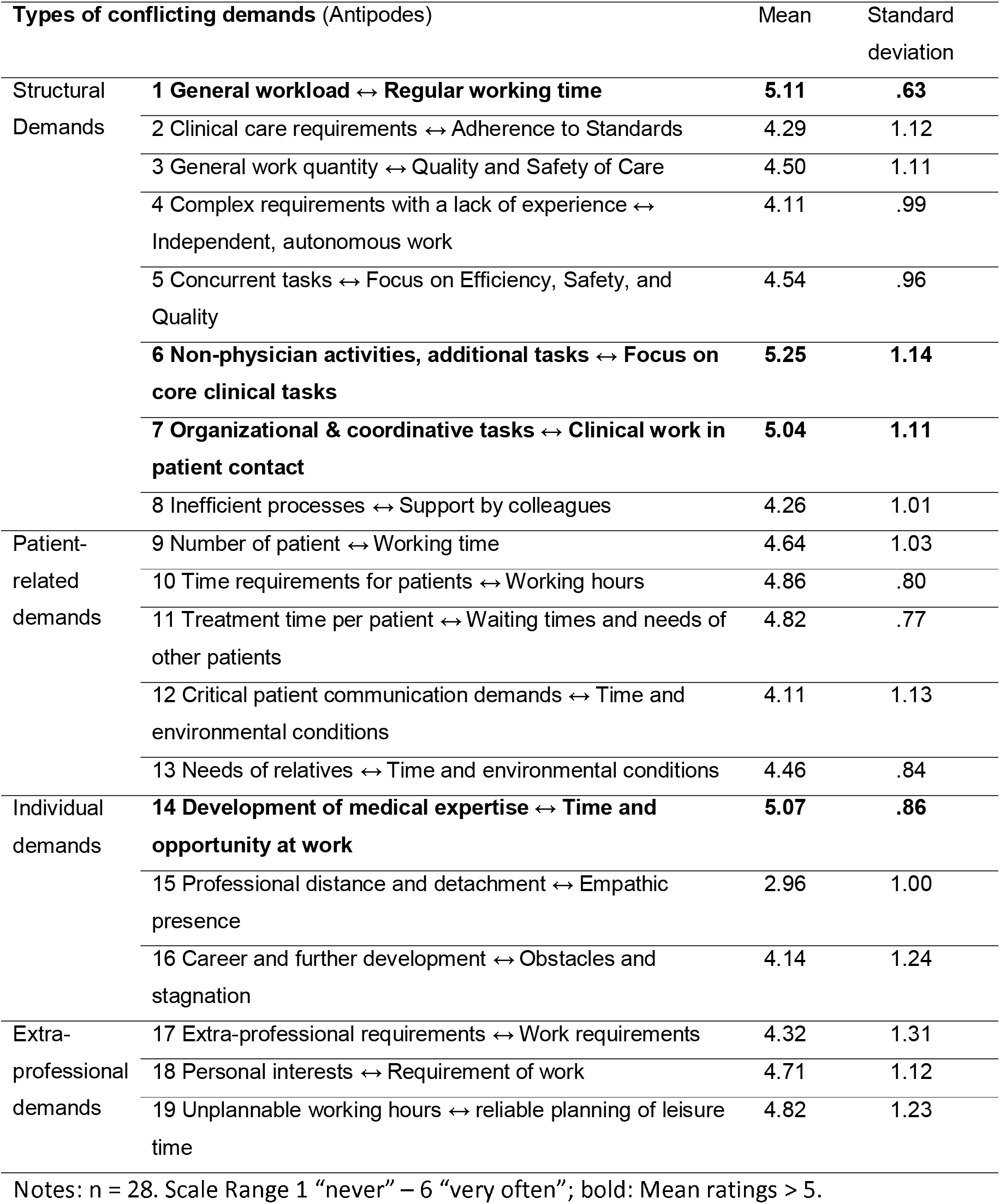
Reported frequencies of conflicting demands among junior physicians.

### For physician well-being on the job outcomes, the following brief measures were applied

Burnout was measured with the German version of the Maslach Burnout Inventory (MBI) with the subscales emotional exhaustion and depersonalisation (21). Within burnout research among physicians, the MBI is the most frequently applied assessment tool (1). Response scale ranged from 1 = never to 6 = very frequent.

We screened for depression with a two-item screening scale (PHQ-2) (22). Responses ranged from 1 = “not at all” to 4 = “almost every day”.

Intention to leave was again assessed with one item (wording: “How often have you thought about quitting your job in the last 12 months?”; scale range: 1 “never” to 5 “every day”).

Job satisfaction was measured with an one item measure (item wording: “How satisfied are you with your job in general?”; scale range: 1 “very satisfied” to 7 “very dissatisfied”).

Additionally, sociodemographic characteristics was collected: Sector of care (inpatient vs. outpatient care), gender (female, male, non-binary), working time (full-/part-time), professional tenure (years of work since approbation), and specialty (free text format being classified into 1 = “specialties with surgical intervention” including participants from anaesthesia, surgery, gynaecology/ obstetrics), 2 = “specialties without surgical intervention” including neurology, cardiology, internal medicine, oncology).

### Analyses

For step 1, interview statements were transcribed verbatim (code book in German language can be obtained from corresponding author). Statements on conflicting demands were independently coded by two study team members, respectively. In case of disagreements, conflicts were discussed and resolved in the study team. Additionally, coders sought to classify each conflicting within one of the pre-defined domains of conflict (see above).

For step 2, we determined descriptive statistics for all 19 items on conflicting demands using SPSS 29.0 (Inc., Chicago, IL, US). For testing of group differences, we used t-tests. For testing associations between physicians’ appraisals on contradictory demands and respective well-being on the job outcomes, we determined Pearson correlation coefficients.

## RESULTS

### (Step 1) Interview results: Exploration of key conflicting demands in junior physician work life

Altogether, seven interviews were conducted with junior physicians undergoing specialty training (3 females, 4 males; mean professional tenure = 2.8 years, Range 2-4 years; 6 full-/1 part-time; 6 inpatient, 1 outpatient care). Interviewees worked across a variety of specialties: 3 internal medicine (intensive care, haemato-oncology, general internal medicine), 2 neurology, 1 ophthalmology, 1 paediatrics.

After content analysis of all statement across the seven exploratory interviews, we identified 19 distinct conflicting demands (for detailed items, see Table 1). Frequency of experiences varied across the four pre-defined categories: the majority of conflicting demands was classified within structural demands (51%); patient-related demands with 20%, individual demands 13%, and 16 in extra-professional demands.

### (Step 2) Survey results on key conflicting demands

Overall, 74 survey forms were sent out. We received completed surveys from 28 junior physicians (response rate: 37.7%). Almost all were employed in a clinic (n=26, 92.9%) with just a minority working in outpatient care (3.6%). Gender was equally distributed (n=14 female, 50%) and, again, the majority was working full time (n=27, 96.4%). The sample stem from over ten different medical specialties, and we classified into surgical, operative specialties (n=10, 35.7%) and medical, conservative specialties (n=17, 60.7%). Mean professional tenure was 3.70 years (SD=2.69, Range 0.1-11 years).

### Concerning the mean ratings of the conflicting task demands, we obtained the following ratings (cf., Table 1)

Across the 19 different types of conflicting demands, we observed almost consistently high frequency ratings. The four highest values, with mean ratings higher than five (i.e., scale levels 5 = “rather often” and 6 “very often”) were the following contradictory task demands (in descending order): ‘Non-physician activities, additional tasks versus Focus on core clinical tasks’ (item #6); ‘General workload versus Regular working time’ (item #1); ‘Development of medical expertise versus Time and opportunity at work’ (item #14); and ‘Organizational & coordinative tasks versus Clinical work in patient contact’ (item #7).

Concerning these four types of conflicting demands, we did not observe differences for reported gender and our overall specialty categories. Concerning professional tenure, we just observed one significant association: higher tenure was associated with fewer experiences of the contradictory demand “Development of medical expertise vs. Time and opportunity at work” (r = -.39, p = .04).

### Physician health and well-being outcomes

Concerning the affective and cognitive-affective outcomes health and well-being outcomes of interest, we obtained the following results:

Altogether, the reported means for our five indicators of junior physicians’ well-being on the job were in a medium range (cf., Table 2).

**Table 2:** Reported physician well-being on the job outcomes.

| Physician outcomes | Scale Range | Mean | Standard deviation |
| --- | --- | --- | --- |
| Burnout – Emotional Exhaustion (Scale mean) | 1 'never' – 6 'very frequent' | 3.86 | 1.13 |
| Burnout – Depersonalisation (Scale mean) |  | 2.40 | 0.91 |
| Depression (Mean sum score, PHQ 2) | 1 'none' - 4 'almost every day' | 3.54 | 1.20 |
| Intention to leave job during past 12 months | 1 'never' – 5 'every day' | 1.96 | 1.02 |
| Job satisfaction | 1 'very satisfied', '7 very unsatisfied' | 2.86 | 1.41 |

Note: n=29

### Associations between frequent conflicting demands and physician outcomes

We then determined associations between the four most frequent conflicting demands and the physician well-being at work outcomes. Table 3 presents the correlational results, respectively:

**Table 3:** Associations between conflicting demands and physician well-being outcomes.

| Conflicting demands | Physician well-being outcomes |  |  |  |  |
| --- | --- | --- | --- | --- | --- |
|  | Burnout - Emotional exhaustion | Burnout – Depersonalisation | Depression | Intention to leave | Job satisfaction |
| 6 Non-physician activities, additional tasks ↔ Focus on core clinical tasks | -,10 | ,17 | ,03 | ,17 | -,09 |
| 1 General workload ↔ Regular working time | ,25 | ,12 | ,02 | ,01 | ,02 |
| 7 Organizational & coordinative tasks ↔ Clinical work in patient contact | ,24 | ,32 | ,18 | -,04 | ,29 |
| 14 Development of medical expertise ↔ Time and opportunity at work | -,34 | ,05 | -,24 | -,27 | **-,40** |

Notes: n=28; bold if p < .05.

Overall, observed associations between the four conflicting demands and the physicians well-being outcomes were weak. We only observed one significant association: higher frequency for conflicts between “Development of medical expertise vs. Time and opportunity at work” was associated with lower job satisfaction (r = -.40, p= .03).

Within additional analyses to test for the robustness of our results, we ran partial correlations with adjusting for sex, specialty domain, and job tenure, respectively. We above reported results did not change substantially after controlling for the confounders (results not reported but can be obtained from the corresponding author).

## DISCUSSION

The first years in clinical work are particularly stressful for junior physicians - especially during course of their post-graduate specialty training. High levels of work stress have been attributed to adverse work conditions during this early stage of the professional career, encompassing a broad spectrum of demands inside and outside of the work environment. This two-step, exploratory study therefore sought to identify experiences of contradictory task demands among junior physicians and to investigate potential associations with self-reports of mental well-being on the job outcomes. We deem the following insights and contributions to the literature base on junior physician work-life factors to be relevant:

First, our investigation sheds lights on contradictory task demands in clinical work, particularly among junior physicians. Junior physicians often face multiple, concurrent challenges in balancing clinical duties, administrative tasks, educational requirements, and personal time (17, 23). This can lead to experiences that incompatible demands imposed by the organization and work environment do not match with available resources, i.e., time, capabilities, etc. (19). Investigations are necessary that deepen our understanding on these specific sources of work stress. They also offer valuable insights into the various contradictory demands that resident physicians face and the impact these demands have.

Using a qualitative and quantitative approach, we identified and classified a wide spectrum of conflicting demands, reported by junior physicians during their first years of clinical work. Eventually, 19 different types of contradictory demands were proposed and surveyed a sample of junior physicians: the observed data highlights the pervasive nature of conflicting demands within the clinical setting, emphasizing the considerable frequency with which these stressors occur in junior physicians’ everyday work life. The four highest-rated conflicts, all with mean ratings exceeding five (scale label ‘rather often’), underline the significant tension between core clinical duties and additional non-clinical responsibilities (17, 19, 23). For instance, the highest-rated conflict ‘Non-physician activities versus Focus on core clinical tasks’ illustrates the strain clinicians face when administrative or ancillary tasks encroach upon their primary patient care duties. Similarly, the conflict between ‘General workload vs. Regular working time’ indicates that clinicians frequently struggle to balance their heavy workloads with the constraints of their scheduled working hours. The third most frequent conflict, ‘Development of medical expertise vs. Time and opportunity at work,’ suggests that clinicians often find it challenging to pursue professional development amidst their demanding schedules. Lastly, the tension between ‘Organizational & coordinative tasks versus Clinical work in patient contact’ further emphasizes the ongoing struggle to balance direct patient care with the myriad of other organizational responsibilities. Overall, frequency appraisals were fairly high with only one item being evaluated below the scale’s mean (item #15: Professional distance and detachment versus Empathic presence).

These findings underscore the complexity of the clinical work environment, where multiple conflicting demands create a challenging landscape for healthcare professionals. Therefore, we sought to explore potential association of experiences of contradictory tasks and well-being outcomes on the job. Across the multiple physicians well-being outcomes, the majority of associations was not significant (i.e., we just observed one significant relationship with more reported dissatisfaction with higher experiences of ‘Development of medical expertise versus Time and opportunity at work’, item #14). Notwithstanding the limited sample size and statistical power, this finding spurs further consideration. Post-hoc, several explanations may apply: first, junior physicians need to cope with a complex array of adverse and challenging work conditions within the clinical learning environment that influence mental well-being on the job (23). Merely focusing on contradictory task demands may neglect other critical demands and resources in the process of work stress (9). Secondly, it has been proposed that certain stressors might be perceived as challenging and legitimate in the context of high-stress and intensive work environments, i.e., undergoing phases of demands that exceed the demands during post-graduate training might be perceived as part of medical training and education and for competence building (24). Thirdly, we did not capture many context factors in course of training that might buffer high loads of contradictory demands, e.g., opportunities for competence development, valuable social support from senior physicians or faculty that might alleviate work stress (24, 25).

### Limitations

Our solely self-reported, cross-sectional findings should be interpreted in the light of several limitations. Although we applied a combination of qualitative and quantitative methods, the studies should be conceived as a first and exploratory approach to enable in the future a deeper understanding on experiences of contradictory task demands among physicians undergoing post-graduate specialty training. External validity should be interpreted carefully since we interviewed and surveyed a small sample of physicians in Germany. The categories and types of contradictory demands established within and from the content analyses need further refinement, cross-validation, and confirmation. Our quantitative analyses using a convenience sample were run without prior determination of statistical power and we did not adjust for multiple testing. We acknowledge that contradictory tasks demands may be different in certain specialties, such that we assume that junior physicians working in acute care environments (e.g., EDs, ICUs, Surgery) might reported different experiences.

### Implications for research and clinical practice

Addressing these conflicts is crucial for improving job satisfaction, reducing burnout, and enhancing the overall quality of patient care. Therefore, targeted interventions aimed at reducing these conflicts could play a vital role in supporting healthcare professionals and ensuring the sustainability of high-quality clinical care. Concerning clinical practice, to the best of our knowledge, there are just a few interventions studies that explicitly addressed conflicting demands in physicians (26)

Concerning future research, further exploration should seek to further understand experiences of contradictory demands and how different goals may interfere or even mutually facilitate learning and skill development, i.e., experiences of overcoming challenges of incompatible demands may help to develop respective coping strategies or alternate future behaviours in similar situation (27). Moreover, there might also be positive ramifications of goal and task conflicts such that goals and means to achieve these become more salient or early considerations of goals conflicts may spur saving or protection of individually valued resources. Lastly, since we merely used self-reports, future studies should strive to explore associations of experiences of conflicting demands in physicians’ work with physiological stress markers, ideally with longitudinal data (28, 29). Lastly, we acknowledge that conceptual overlap with other work stress sources need to be carefully considered: for instance, similarities with other concepts such as role conflicts in junior physicians (17) or work-life (in)balance (23).

## Conclusion

Junior physicians often struggle to balance their everyday workload with educational requirements and patient care. Especially if demands exceed available resources, junior physicians need to prioritize tasks under time constraints, often facing contradictory demands that force them to make difficult decisions about where to allocate their time and attention. Our exploratory survey sought to capture experiences of contracting demands among junior physicians undergoing specialty training. We established and surveyed a list of contradictory demands and tested for associations with well-being outcomes on the job. Our work provides base for future investigations into work-life complexities of junior physicians, including conflicting demands, and the coping strategies they employ to manage these challenges.

## Data Availability

All data produced in the present study are available upon reasonable request to the corresponding authors (MW).

## Acknowledgements

This work has been part of the Doctoral Thesis Requirements for Ms. Susann Boehm (Medical Faculty, LMU Munich). We gratefully acknowledge both time and expertise of contributing participants.

